# Disease Triangle Dynamics of Coccidioidomycosis in Northern Arizona

**DOI:** 10.1101/2021.06.24.21258773

**Authors:** Heather L. Mead, Dan Kollath, Marcus de Melo Teixeira, Chandler C. Roe, Paul K. Keim, Carmel Plude, Joel Terriquez, Nivedita Nandurkar, Chelsea Donohoo, Brettania L.W. O’Connor, Bridget M. Barker

**Affiliations:** Pathogen and Microbiome Institute, Northern Arizona University, Flagstaff, AZ. 86011. U.S.A; Translational Genomics Research Institute (TGen), Phoenix and Flagstaff, AZ. USA; Faculty of Medicine, University of Brasilia, Brasilia. Brazil; Northern Arizona Healthcare, Flagstaff, AZ. 86001. U.S.A; Coconino County Health Department, Flagstaff, AZ. 86004. U.S.A; Department of Health Sciences Northern Arizona University, Flagstaff, AZ. 86011. U.S.A

**Keywords:** coccidioidomycosis, Valley fever, disease triangle, fungal infection, diseases of southwestern united states, endemic fungi, Northern Arizona

## Abstract

*Coccidioides* species are the etiological agent of Valley fever (Vf). Disease ranges from pneumonia to extrapulmonary infection. If diagnosis is delayed or missed, the risk of severe disease increases. We investigate the disease triangle (intersection of pathogen, host, and environment) of Vf in Northern Arizona, where the risk of acquiring the disease is lower but prevalent and rising. We inspect reported and hospitalized cases of VF hosts. Then assess pathogen origin by comparing Northern Arizona clinical isolates to isolates from other regions. Lastly, we survey Northern Arizona soils for evidence of *Coccidioides*. We found that cases of Vf are increasing some requiring hospitalization. The Northern Arizona *Coccidioidies* isolates were genetically related to Southern Arizona populations. However, we detected *Coccidioides* DNA in Northern Arizona soils. We expect that cases will continue to increase and suggest increased awareness and screening for the disease is crucial to limit severe illness in Northern Arizona.

**Article Summary Line:** Our work is the first description of the Valley fever disease triangle in Northern Arizona, which considers the complex interaction between host, pathogen and environment in the region. Our data suggests that the prevalence of diagnosed cases is rising in the region, some of which are extreme enough to necessitate hospitalization. We present the first evidence of *Coccidioides* spp. in Northern Arizona soils, suggesting that the pathogen is maintained in the local environment. Until disease prevention is an achievable option via vaccination, we anticipate that incidence of Valley fever will rise in the area. Therefore, enhanced disease awareness and screening for the coccidioidomycosis is vital to the communities of Northern Arizona.

## Introduction

The disease triangle is a concept which includes the interaction between host, pathogen and the environment. Infectious diseases can emerge when a vulnerable host, virulent pathogen and favorable environmental conditions complete the triangle (1). This framework has been applied to a wide variety of diseases that affect both plants and animals (2, 3). Lessons can be learned from the famous potato famine in Ireland and applied to protect future crop security (1). Similarly, during the global COVID-19 pandemic these interactions are under scrutiny to determine and monitor the animal reservoir of the novel SARS-CoV-2 virus (4-6). Clearly, the disease triangle is a critical perspective for considering infectious diseases and predicting future implications for human health. One environmentally acquired fungal infection of concern is coccidioidomycosis (Valley fever), caused by two *Coccidioides* species. Alarmingly, cases of this disease are increasing nationwide (7-9).

Valley fever in humans can be self-limited, requiring little to no medical care, or chronic causing years of treatment and/or life-long symptoms (10). Infection occurs when aerosolized environmental arthroconidia are inhaled by a susceptible host, thus, Valley fever typically manifests as a respiratory infection which presents as asymptomatic, acute or fibrocavitary chronic pneumonia (11). In severe cases, dissemination to extrapulmonary sites, such as the skeletal or the central nervous system occurs and requires life-long antifungal treatment (10, 11). This broad range of clinical symptoms makes determining yearly case burden challenging, but estimates range from 150,000 - 350,000 cases/year in the United States (8, 12). Fortunately, for many patients the disease can be asymptomatic or present as self-limited pneumonia not requiring clinical intervention. Unfortunately, for symptomatic patients, misdiagnosis occurs frequently, which contributes to the inappropriate use of antibiotics or long delays in correct treatment (9, 13). Therefore, reported cases underestimate actual cases due to individuals not seeking medical care, misdiagnosis and underreporting (8, 14). In known endemic regions, approximately 30% of community acquired pneumonia cases are known to be due to Valley fever, but patients are infrequently tested for Valley fever and thus not reported (15-17). Preventing infection is difficult because even at individual locations the occurrence of detectable airborne fungal particles can fluctuate daily, consequently any public health warnings based on detection would delayed (18, 19). The underlying mechanisms which contribute to host susceptibility are elusive, preventing the identification of all highly susceptible individuals as a prevention strategy (20). Lastly, despite decades of research, a viable vaccine has not been identified (21, 22). Therefore, an important part of combating this disease currently requires defining the patterns of host susceptibility, pathogen virulence potential and environmental requirements which support the lifecycle. Which approach contributes towards improved awareness among community members, healthcare professionals and infectious disease researchers.

Retrospective studies based on medical records are typically used to identify host specific factors which contribute to the Valley fever disease triangle (16, 17, 23-26). This approach is helpful but incomplete as asymptomatic or mild cases that did not require a medical visit are not included. Nevertheless, these strategies have identified several factors which increase host disease susceptibility. Substantial evidence shows there are differences in risk of Valley fever disease severity by race. Individuals of African or Filipino descent are a recognized risk group for severe disease, in particular disseminated infection (26). Recently, Native American populations have been suggested to have increased dissemination and hospitalization rates (27). The underlying mechanisms responsible for increased disease vulnerability are undefined (28). These observations are complicated by the uneven distribution of races in endemic areas, uneven sample sizes and the potential effect of socioeconomic and other underlying factors. These complications do not nullify the indication of increased risk among specific populations, but rather validate the need for in-depth studies which consider these aspects (29). Lastly, while many fungal diseases are restricted to immunocompromised individuals, coccidioidomycosis occurs in seemingly healthy individuals as well which could be due to undescribed host factors (30-32). In general, many host specific aspects which dictate Valley fever disease outcomes are undefined or incomplete, which prevents identifying all high-risk populations(9, 25).

The evolution of a host-specific lifecycle, which is distinct from the environmental lifecycle, contributes to *Coccidioides’* virulence potential causing disease in humans and other mammals (33). When the environmental fungal arthroconidia (asexual spore structure) are inhaled by a mammal, they swell into large structures called spherules which are uniquely adapted for survival and proliferation in the host (34, 35). These fungi have developed thermotolerant strategies to survive at high temperatures, such as those encountered in the human body, subsequently increasing pathogenic potential (36). In addition, genomic analyses suggest that in comparison to common ancestors *Coccidioides* experienced a reduction in genes needed to degrade plant material, a common nutrition source for many fungi, and gained the ability to degrade a variety of animal proteins (37). Molecular systematics and taxonomical studies have determined that the *Coccidioides* genus is comprised of two species, *C. immitis* and *C. posadasii* (38, 39). Both of these species create the host specific endosporulating spherule structure, that is not observed in other dimorphic fungi (33). Each species has defined subpopulations with substantial genetic variation (40-42). Currently, it is unknown if virulence or the ability to cause disease varies among strains beyond what has been observed in mice or in dated literature (30, 43-46). So far, no specific allelic distributions of fungal virulence phenotypes are associated with specific disease symptoms. This is in part because little work has been conducted in this area, though variation in virulence among isolates of *Coccidioidies* has been suggested (43, 45, 47). This missing and crucial information specific to virulence potential could explain the observed disparity in disease manifestation among individuals and should be investigated.

The environmental niche which supports existence of *Coccidioides* has not been completely defined but it is hypothesized that the fungi co-evolved with desert mammals, such as rodents (48, 49). Thus, the pathogen can frequently be detected in the tissue of wild mammals, burrows, or archeological sites that are likely enriched with animal-derived organic matter (39, 50-53). In these situations, it is suggested that the fungi exist as mammalian endozoans which survive within the host without causing symptoms and benefit by having direct access to nutrition upon host demise (48). The known environmental distribution of the pathogen includes arid zones of Latin America, ranging from Mexico to Argentina and the southwestern United States (40, 54). Recently, the recognized range of the pathogen expanded with locally acquired cases occurring in northwestern states, such as Washington (55, 56). Alarmingly, the environmental distribution of *Coccidioides* is predicted to continue to expand in response to climate variability (57, 58). Regions of the southwest that were not traditionally considered endemic will likely serve as suitable habitat for the pathogen in the near future and subsequently it is expected that cases of Valley fever will continue to rise (59).

Historically, Arizona has reported the highest number of diagnosed cases of Valley fever nationwide each year, except in 2018 when Arizona was surpassed by California (7, 9). In moderately or suspected endemic regions such as Northern Arizona, the risk of acquiring disease is lower than in established endemic regions, but still prevalent (60). Subsequently, infection is often attributed to travel or prior residence in highly endemic regions. Because the risk of Valley fever is lower across Northern Arizona compared to highly endemic counties of southern Arizona there is potential that diagnosis can be delayed or missed due to lack of awareness and testing (61). Northern Arizona encompasses a third of the state and is divided into five counties, Mohave, Yavapai, Coconino, Apache and Navajo (Figure 1). As with the rest of the state, the incidence of new cases in Northern Arizona is on the rise (62, 63). However, data on patients/cases from the region is often grouped with the rest of the state, with most discussion focused on the southern counties Maricopa, Pima and Pinal, with the highest proportion of reported cases. The environmental factors and potential host population demographics in Northern Arizona are very different from counties to the south. The region is predicted to further support the pathogen’s survival in the near future, but little focus has been placed on the current disease burden of the region (58).

**Figure 1.**
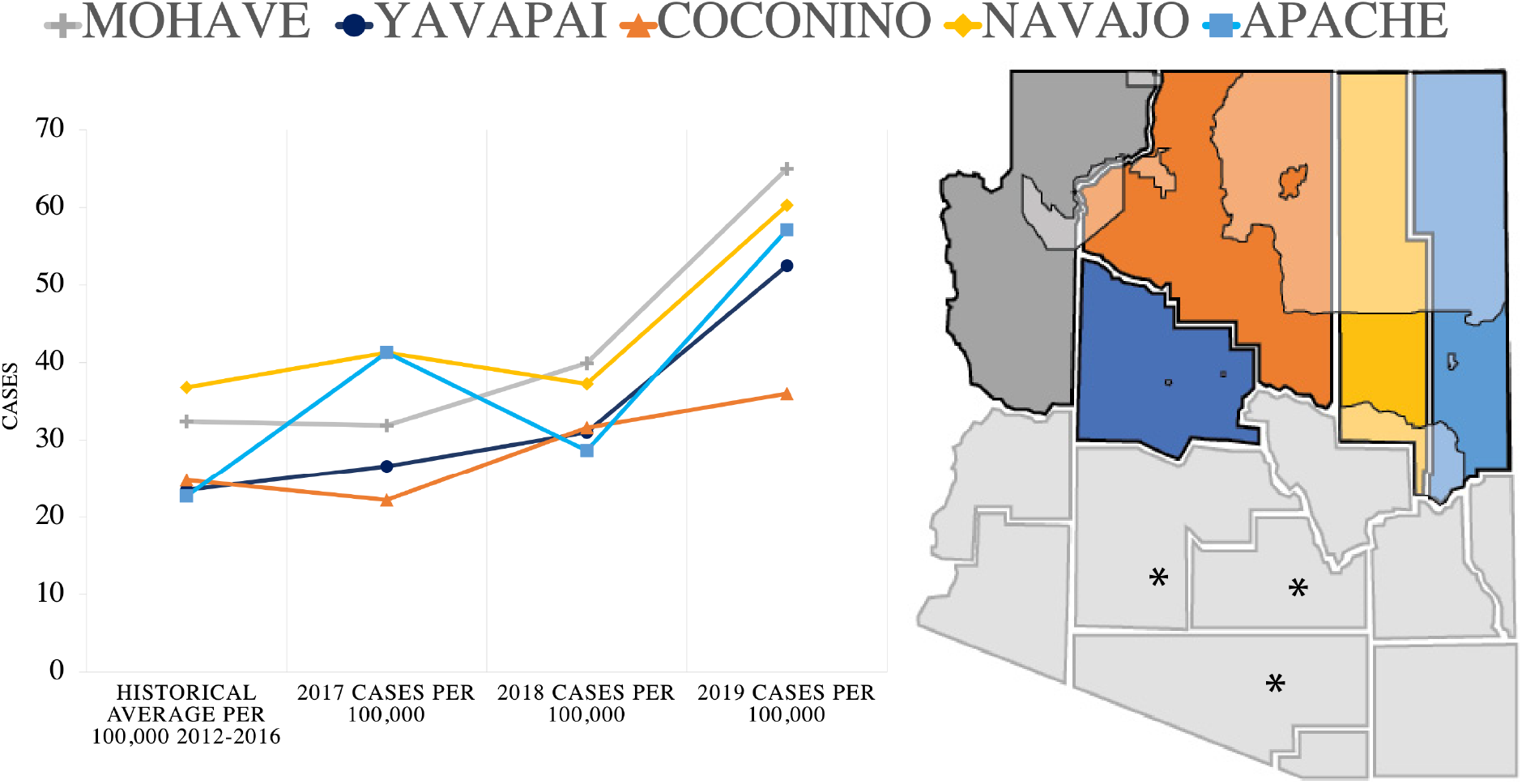
Northern Arizona county location and Valley fever cases. Yearly, the number of cases per 100,000 population are increasing compared to historical averages in all Northern Arizona counties. Geographic location of northern Arizona counties. Left to right, Mohave, Yavapai, Coconino (largest), Navajo and Apache. Tribal land located in Northern Arizona counties are shaded. The southern counties which experience the highest yearly case counts per 100,000 are denoted with asterisks.

Herein, we conduct the first focused investigation of coccidioidomycosis in humans in Northern Arizona. We investigate susceptible populations, origins of infection and the potential for the environment to support the pathogen. Our goal is to document the region-specific implications for human health in regard to Valley fever, providing a reference for future investigations and public health awareness campaigns.

## Methods

This project utilized four data sources. 1) Public health records to capture reported Valley fever in Northern Arizona counties between year-year. 2) Hospital chart data to capture in patient Valley fever trends at Northern Arizona Healthcare 3) Genomic DNA from clinical isolates obtained at the Northern Arizona Healthcare. 4) DNA extracted from soil in Northern Arizona counties.

1. Public Health records: Yearly, county case reports (2017, 2018 and 2019) and historical averages (2012-2016) per 100,000 population were accessed through the Arizona Department of Health Services (AZDHS) website.
2. Hospital Records: A retrospective chart review was performed to identify all in-patients that were screened for Valley fever during an 18-month period (7/1/17-12/31/18) at the Flagstaff location of Northern Arizona Healthcare. Screening was defined as when a patient was subjected to Valley fever evaluations through x-ray, titer or complement fixation serological diagnostics. Confirmed positive diagnoses were defined as; positive IgM, titer >1.8. These confirmed positive cases were reviewed by the infectious disease department to definitively confirm diagnosis. Individual medical records with confirmed coccidioidomycosis cases were further investigated for demographic and comorbidity information.
3. Fungal Isolates: The fungal clinical isolates were previously collected and stored under IRB No. 764034 as part of the Northern Arizona University Biobank study with Northern Arizona Healthcare. For this study, these isolates (Supplemental Table 1) were transferred to the Pathogen and Microbiome Institute biosafety level three (BSL-3) facility. Personal protective equipment and pathogen manipulation occurred as previously described recommended by the Biosafety in Microbiology or Biomedical Laboratories (64, 65). Fungal Growth, Nucleic Acid Extraction and Sequencing: Fungal isolates were cultured as previously described (65, 66). Briefly, to confirm pure cultures the isolates were grown on 2xGYE (2% glucose, 1% yeast extract agar) at 30°C, for seven days. Next, small plugs of each isolates were used to inoculate liquid 2xGYE (2% glucose, 1% yeast extract) in vented baffled Erlenmeyer flasks and grown at 30°C, for six days on a shaking incubator. These cultures were heat inactivated at 80°C for 30 min in fungal lysis buffer. Genomic DNA was obtain used traditional phenol:choloroform solvents and precipitated using isopropanol and EtOH. DNA quantity and quality was visualized on a 1% agarose gel. These samples were submitted for sequencing at the Translation Genomics Research Institute on a MiSeq V3 kit for 600 cycles. Raw reads have been deposited under accession number PRJNA722304. Population *Analysis:* Raw reads for 60 previously published *C. posadasii* samples representing the Caribbean, Arizona, Texas/Mexico/South America clades (40) and the seven new clinical isolates were aligned to reference *C. posadasii* stain Silveira genome, accession number JADMCF000000000. Using the Northern Arizona Sequencing Pipeline (67) in order to retrieve DNA mutations. The resulting SNP (Single Nucleotide Polymorphism) matrix was used to build a Maximum Likelihood tree using IQ-TREE version 1.6.1, based in 152,663 parsimony informative SNP’s (68). The phylogenetic tree distribution was visualized using Figtree v1.4.4 (69).
4. Soil Collection, Nucleic Acid Extraction and Amplification: A total of 171 soil samples collected between 2018-2020 were included in this study (see Supplemental Table 2 for breakdown of samples and site descriptions). The sites in Northern Arizona drastically vary in ecosystem characteristics from each other as well as from habitat in the highly endemic Southern Arizona region for Valley Fever. Soils were collected and processed as previously described (49). Briefly, each sample was collected with a sterile garden trowel and stored in sterile 50 mL collection containers at room temperature until processing. DNA was extracted following kit recommendations using the Qiagen DNeasy Powersoil Pro Kit (Qiagen, Boston, MA, USA). The presence or absence of *Coccidioides* spp. DNA in soil samples was determined by two highly specific Taqman-based CocciCDX and CocciENV real-time qPCR assays using a Quant Studio 12k Flex Real-Time PCR System (ThermoFisher Scientific (70, 71). Each 20uL reaction contained 1 x Taqman Universal Master mix II (ThermoFisher Scientific), PRIMERS, with DNA template with the following thermocycling conditions: initial denaturation for 10 min at 95°C followed by 40 cycles of 15 s at 95°C and 1 min at 60°C. Three technical replicates were run for each DNA sample. A reaction was considered positive if it showed logarithmic amplification, produced a CT value of <40.

## Results

We sought to identify the number of recent diagnosed cases (2017-2019) of coccidioidomycosis in Northern Arizona compared to historical averages (2012-2016) and to compare the host specific factors of these cases to statewide trends. After which we focused on the in-patient Valley fever patient population of Northern Arizona by performing a chart review during a similar (18 month) time frame at the regional hospital. We then investigated the genetic structure of fungal isolates collected and stored at the hospital compared to the recognized populations of *Coccidioides*. Lastly, we searched for evidence of pathogen presence in Northern Arizona soils, completing our epidemiologic triangle approach.

### Diagnosed Valley fever cases are increasing in Northern Arizona counties

Northern Arizona counties are experiencing increased cases of Valley fever per 100,000 population compared to the historical trends defined by the health department (Figure 1). We focused on 2017-18 as this time frame coincides with the hospital data available to us and chose to also include 2019 data. In Mohave, Yavapai and Apache counties the number of cases per 100,000 population doubled by 2019 in comparison to historical averages (Figure 1, supplemental table 3). These findings are similar in counties across the state. The greatest number of cases reported each year occur in Maricopa, Pima and Pinal counties which are located in Southern Arizona (Figure 1). In general, all of the counties in Arizona are reporting an increase in diagnosed cases each year and an increase in incidence per 100,000 population (supplemental table 3 and 4) with a few exceptions. The age and sex of individuals testing positive for Valley fever in northern counties are similar to statewide observations (supplemental table 5).

### Valley fever hospitalizations in Coconino county

Next, we surveyed all in-patient records at the regional hospital, Northern Arizona Healthcare (NAH) for an 18-month period (7/13/2017-12/31/2018). The relatively short timeframe of this retrospective review is a limitation of this study. Therefore, the previously mentioned county case data was identified to complement <u>the</u> timeframe of this chart review. This regional medical center is located in Coconino county and often accepts patients from other Northern Arizona counties. During the time frame we investigated, there were 124 in-patients screened for Valley fever, 38 of them were positive for coccidioidomycosis (IgM 28, titer 16). Because statewide coccidioidomycosis hospitalization demographic data is not publicly available we compared these in-patient cases to all reported coccidioidomycosis case in Arizona in respect to sex and age (supplemental table 5, Figure 2A). In Arizona, reported cases of Valley fever is distributed approximately 50/50 among males and females with slight fluctuation each year (60). Of the 38 hospitalized patients there were 23 males (60.5%) and 15 females (39.5%). The greatest proportion of Arizona confirmed cases and in-patient cases occurred among individuals between 45-64 years of age. Among all 38 Valley fever hospital patients, we observed a range of comorbidities; diabetes (16%), cancer (10%), HIV (5%) or in some cases multiple conditions (25%). In total 23% of hospitalized patients did not have notable co-morbidities and were otherwise healthy (Figure 2B). For the 38 hospitalized cases, 82% of the fungal infections were restricted to pulmonary locations, and 18% experienced dissemination to other regions of the body (Figure 2C). Nationally, dissemination only occurs in between 1-5% of reported Valley fever cases (25).

**Figure 2.**
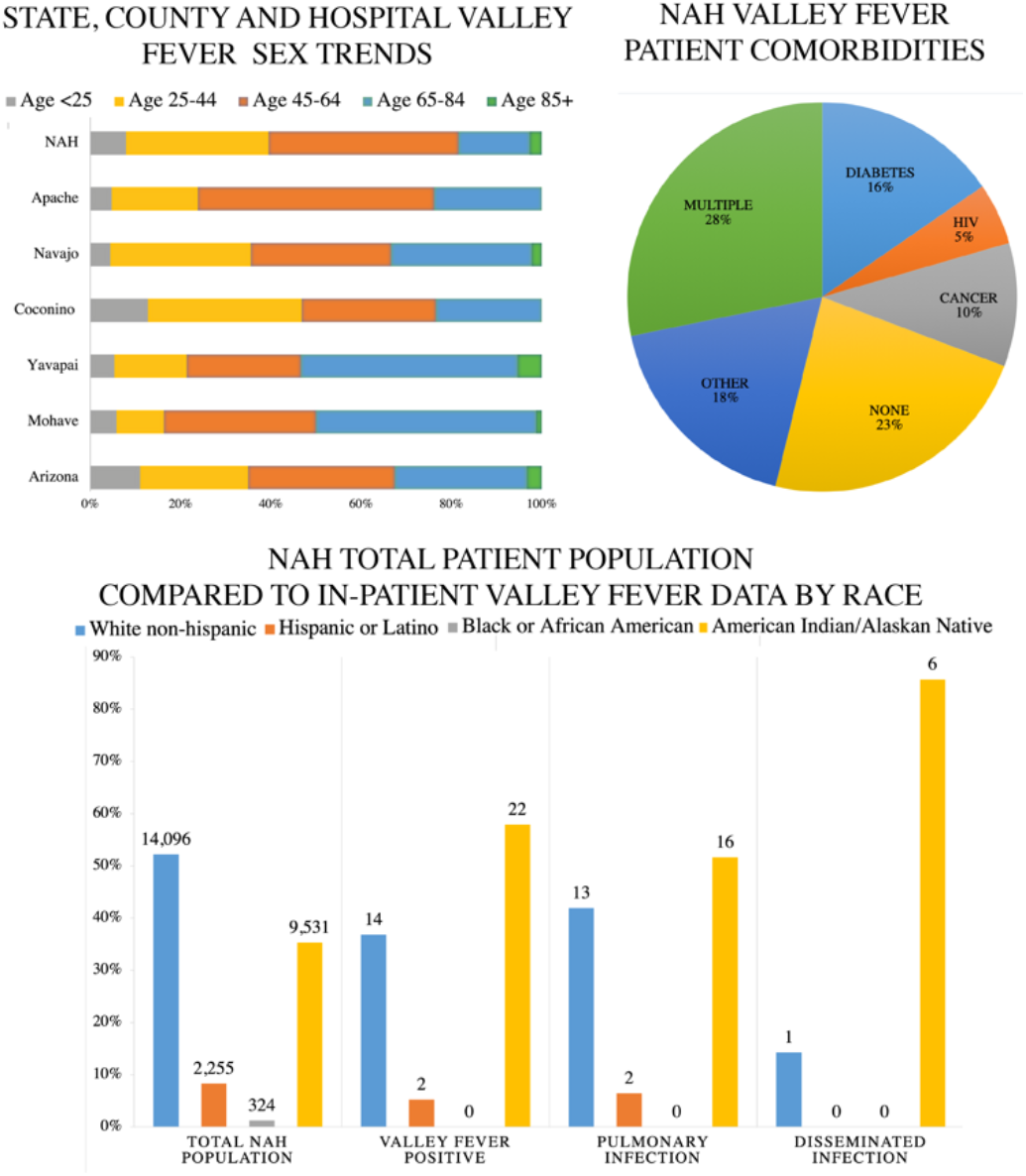
Northern Arizona Valley fever data demographics. 2A) Age of patients diagnosed with Valley fever in Arizona during 2018, Northern Arizona counties during 2018, and regional hospital (NAH) between (7/13/2017-12/31/2018). 2B) Among hospitalized Valley fever patients at NAH 23% of patients had no documented pre-existing conditions. The other 77% had existing co-morbidities such as diabetes (16%), HIV (9%), cancer (10%), and is several cases multiple immunocompromising conditions. 2C) The total population treated at NAH is displayed next to in-patient Valley fever cases for reference. Out of the 38 patients diagnosed with Valley fever from (7/13/2017-12/31/2018) pulmonary infection occurred in 31 (82%) cases, dissemination to extrapulmonary locations occurred in 7 (12%) of the cases. There were 14 white (36.8%), 2 Hispanic (5.2%) and 22 Native American/Alaskan Indian (57.8%) individuals.

**Figure 3.**
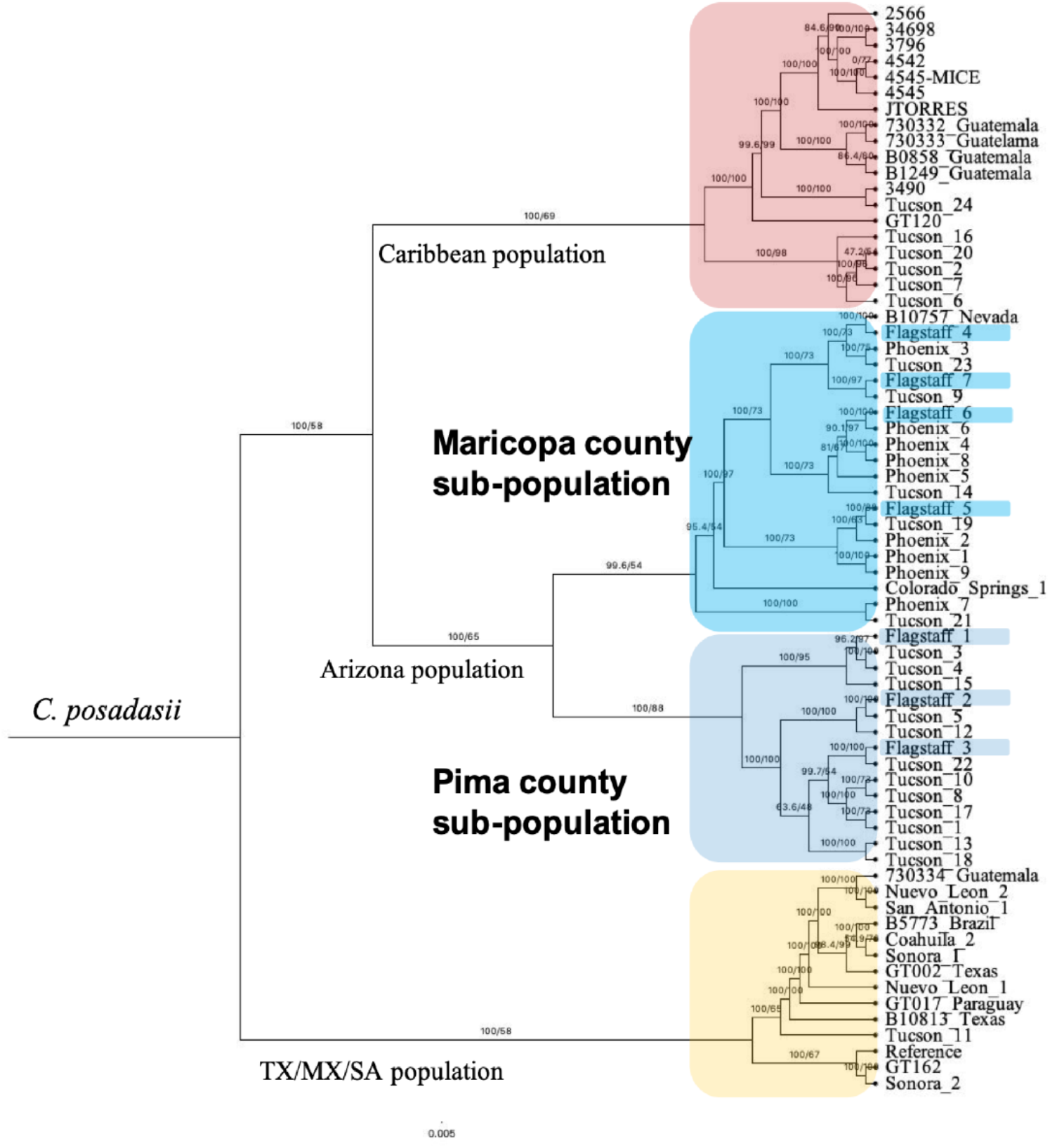
Relationship of Northern Arizona Clinical isolates within *C. posadasii* population suggests traveled related infection. The seven new clinical isolates (highlighted) group with previously published isolates recovered from patients in Maricopa or Pima county populations. *C. posadasii* is comprised of geographically distinct populations designated as Caribbean (top, red), Texas/Mexico and South America (bottom, yellow) and Arizona (middle, blues). Subpopulation structure in Arizona suggests genetically distinct groups with Arizona (Maricopa county; Phoenix and Pima county; Tucson). Mid-point Rooted Maximum Likelihood Tree built using 61 previously published and seven new isolates.

Next, we sought to compare racial demographics in NAH Valley fever cases with Arizona trends. Published literature suggests that individuals with African American or Filipino backgrounds are at increased risk for severe Valley fever (26, 72). Shockingly, in Arizona race/ethnicity is documented for only ∼29% of diagnosed cases (Table 1). This makes assessing especially vulnerable populations based on racial variation challenging and potentially inaccurate. At NAH, we were able to identify race/ethnicity in all cases (Table 1, Figure 2C).

**Table 1.**
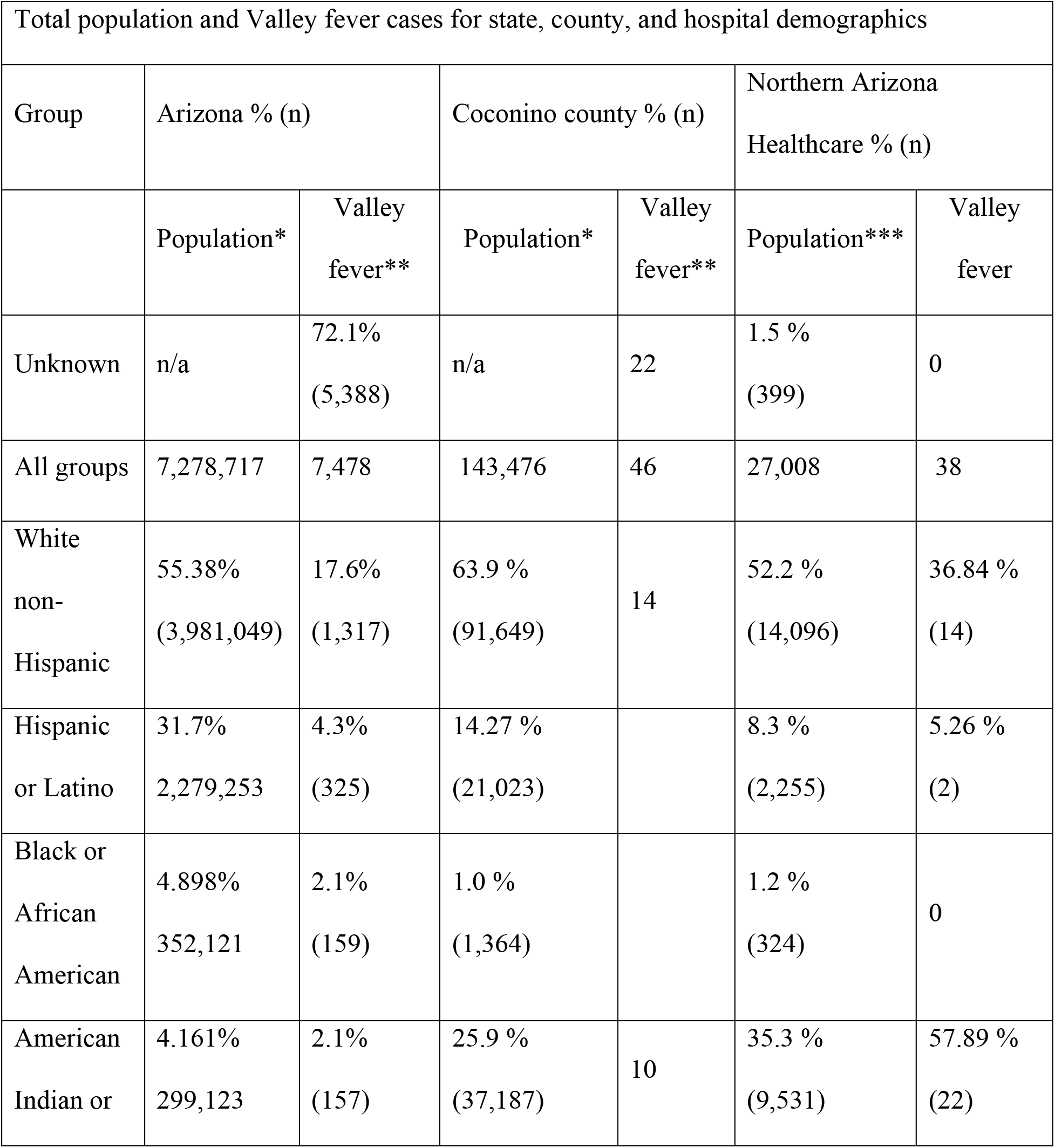
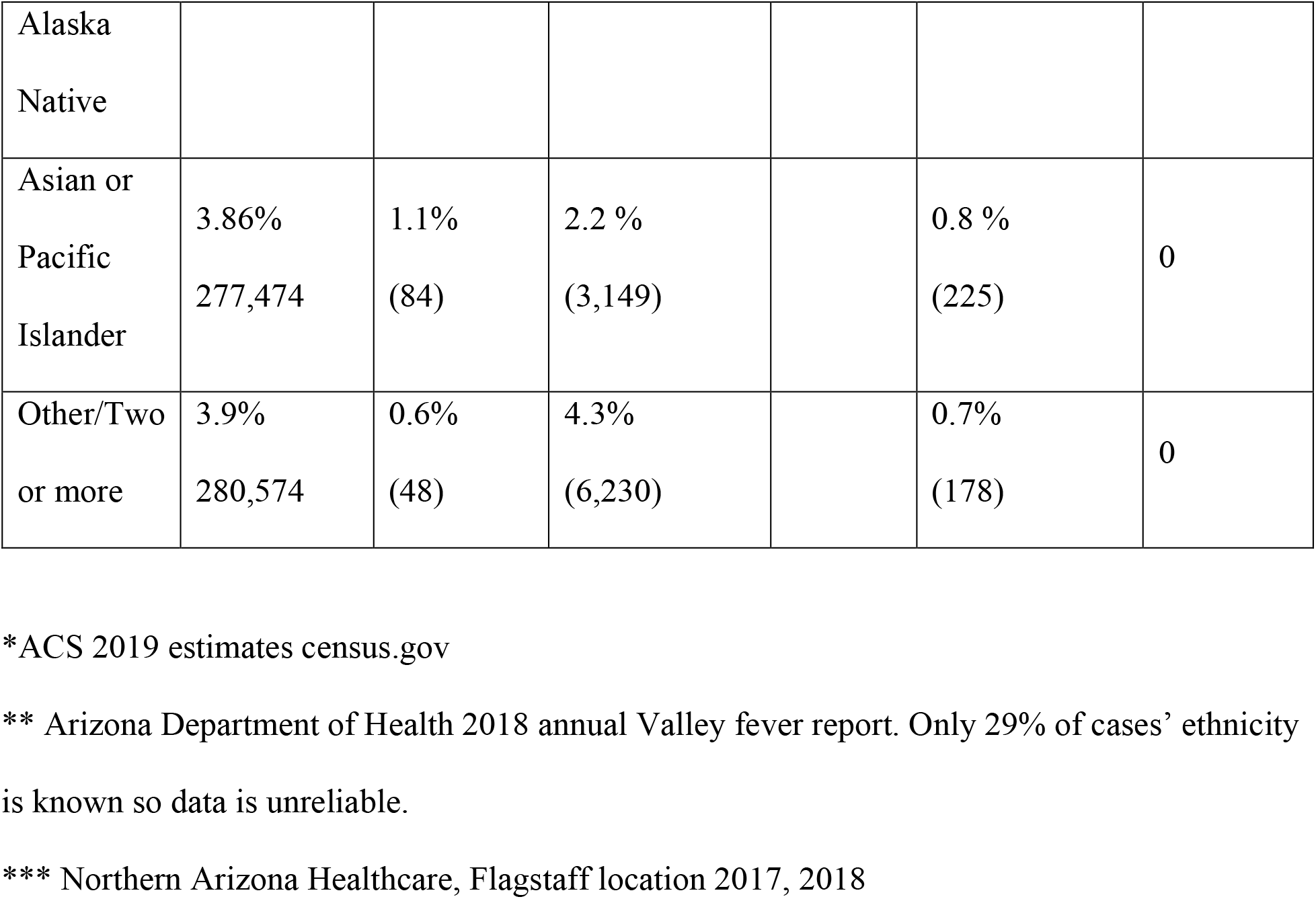
Breakdown of unknown and known race distribution for Valley fever cases in Arizona, Coconino county and Northern Arizona Health Care.

There were 14 white (36.8%), 2 Hispanic (5.2%) and 22 Native American/Alaskan Indian (57.8%) cases. The observed proportion of diagnosed cases among individuals with Native American/Alaskan Indian heritage appears high (57.8%) despite the population prevalence (25.9%) in Coconino county and at NAH (35%) compared to other ethnic groups (Table 1). This trend may be similar to findings released by the CDC which documented increased hospitalization in native American populations with Valley fever. However, statewide examining data when race is known, only 2.1% of the cases are observed in Native American/Alaskan Indian individuals, who make up 4.1% of the state’s total population (Table 1).

### Northern Arizona Clinical isolates originate from Arizona Coccidioides populations

We obtained seven *Coccidioides* isolates from the Northern Arizona Healthcare patient population. We used 60 previously published whole genome paired end reads and recently release PacBIO assembly, *C. posadasii* strain Silveira reference to build a SNP (single nucleotide polymorphism) based matrix. A total of 82.72% or 23,336,819 bases mapped to the reference genome from which we detected 270,853 SNP’s. Next, the IQTREE software was used to identify the best nucleotide substitution model, TVM+F+ASC+R5 using 1,000 bootstrap iterations to the distinguish the Maximum Likelihood tree. This analysis demonstrated that the isolates obtained from patients at Northern Arizona Healthcare are genetically similar to either isolates from Maricopa (n=4) or Pima (n=3 counties) and no unique genetic clusters were found for the Northern Arizona samples.

### Coccidioides spp. are present in Northern Arizona soil samples

We surveyed 171 soil samples obtained between 2018-2020 from Mohave, Yavapai, Coconino, Navajo and Apache county for the evidence of *Coccidioides* DNA. We identified positive samples in all of the counties, with varying rates of positivity (Figure 4, Table 2). In Mohave county there were 8/24 positive soil samples using both PCR assays (CDx and CocciEnv). In Yavapai county there were 6/18 using CDx and 4/18 based on CocciENV results. In Coconino county there were 6/75 positive CDx soils and none with CocciENV. Navajo county had 2/32 CDx positive and none with CocciENV. Lastly, Apache county 2/22 soils which were CDx positive and 3/22 CocciENV positive. In summary, 39 samples were positive for at least one assay (Table 3). It is common to have varying results between the two PCR methods, both which target *Coccidioides* specific alleles. Therefore, our results suggest that *Coccidioides* species are present in Northern Arizona soil. If we consider samples positive for both PCR methods, we identified six positive soils, which provides evidence that *C. posadasii* is present at these sites.

**Figure 4.**
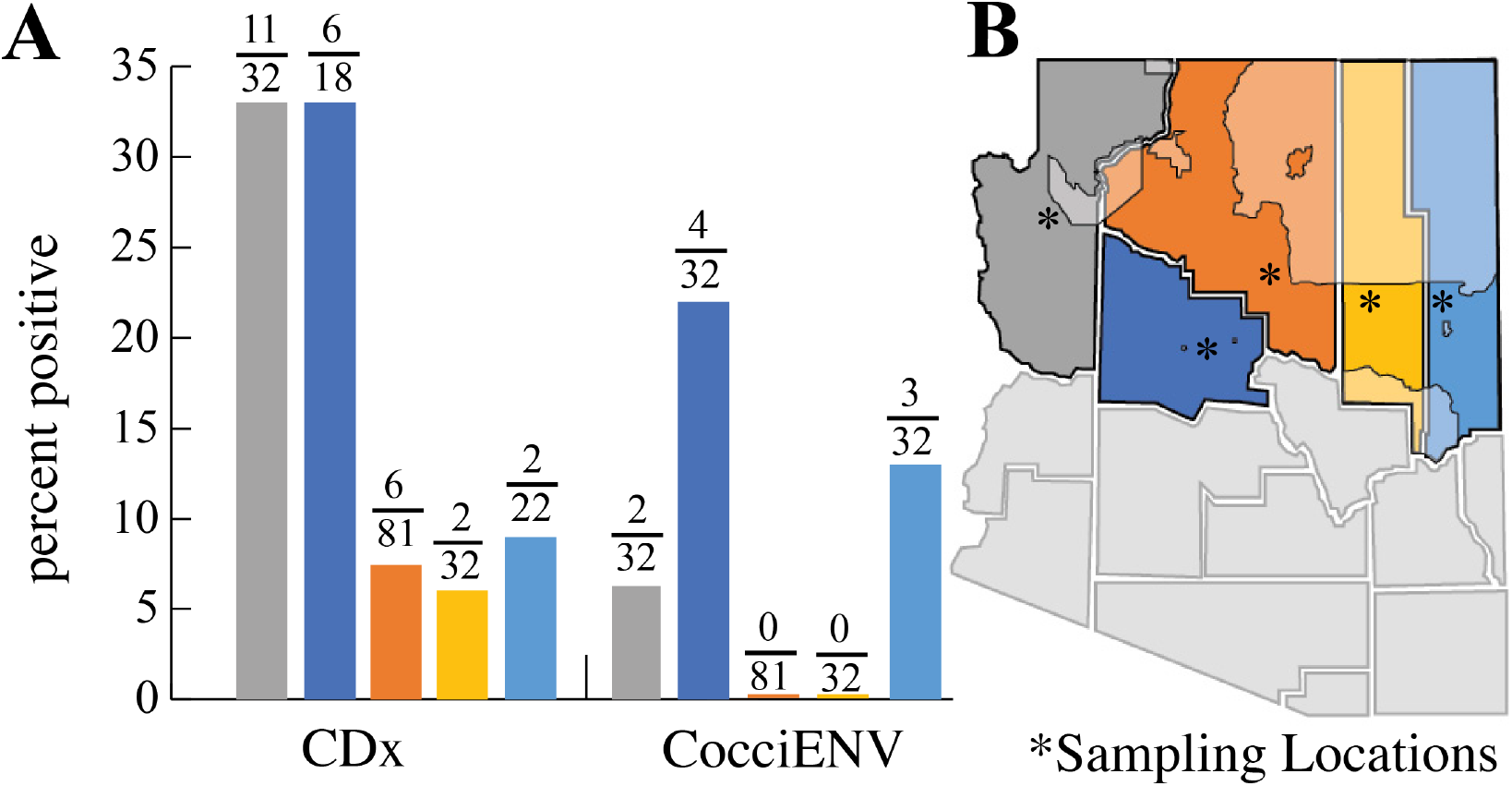
Northern Arizona soils are positive for *Coccidioides* DNA. 4A) Percent positivity for soil in Mohave, Yavapai, Coconino, Navajo and Apache county. 4B) Approximate location of soil collection sites designated with asterisk *. Tribal land located in Northern Arizona counties are shaded.

**Table 2.**
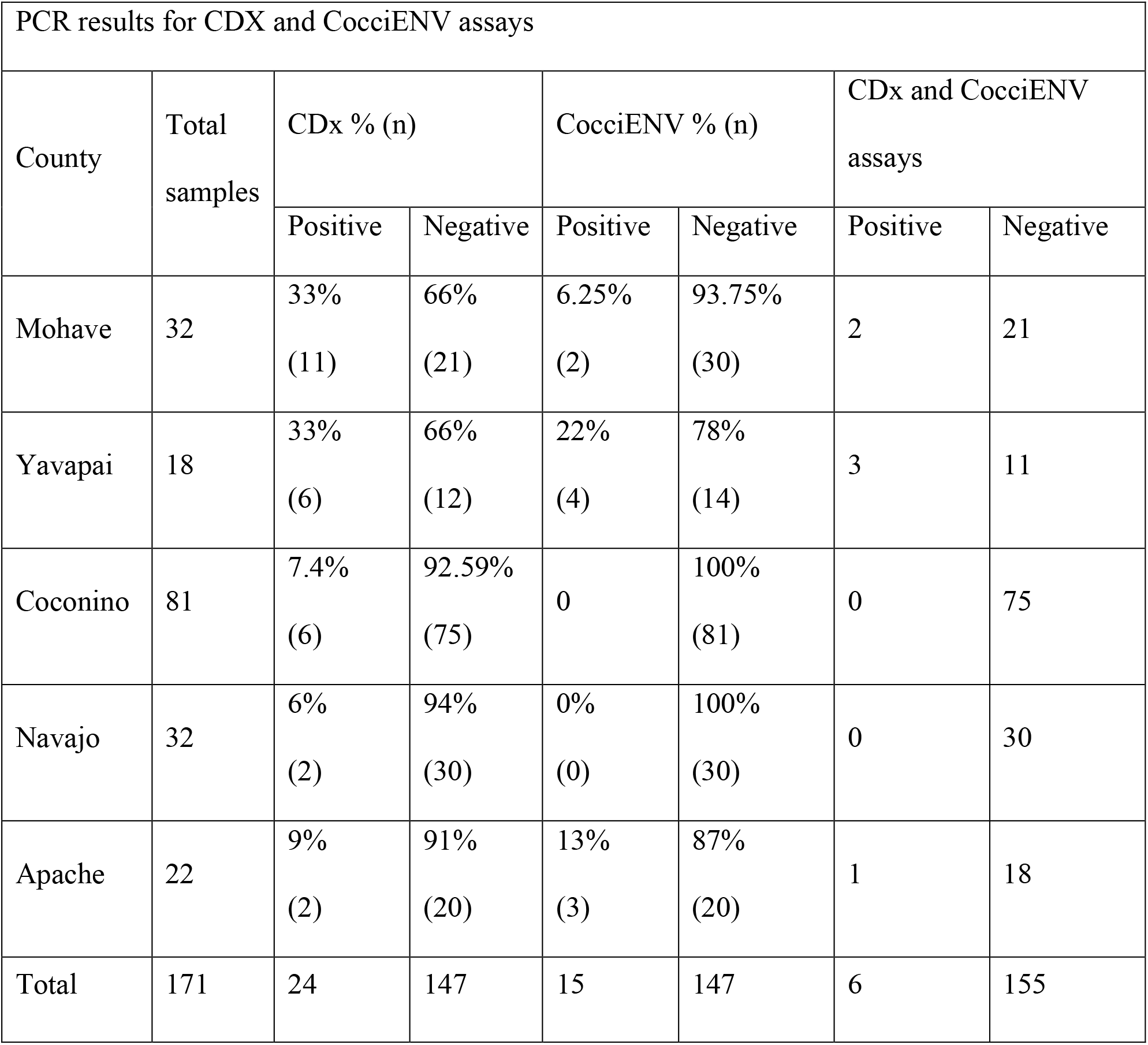
Evidence for *Coccidioides* in northern Arizona county soils.

| PCR results for CDX and CocciENV assays |  |  |  |  |  |  |  |
| --- | --- | --- | --- | --- | --- | --- | --- |
| County | Total samples | CDx % (n) |  | CocciENV % (n) |  | CDx and CocciENV assays |  |
|  |  | Positive | Negative | Positive | Negative | Positive | Negative |
| Mohave | 32 | 33%<br>(11) | 66%<br>(21) | 6.25%<br>(2) | 93.75%<br>(30) | 2 | 21 |
| Yavapai | 18 | 33%<br>(6) | 66%<br>(12) | 22%<br>(4) | 78%<br>(14) | 3 | 11 |
| Coconino | 81 | 7.4%<br>(6) | 92.59%<br>(75) | 0 | 100%<br>(81) | 0 | 75 |
| Navajo | 32 | 6%<br>(2) | 94%<br>(30) | 0%<br>(0) | 100%<br>(30) | 0 | 30 |
| Apache | 22 | 9%<br>(2) | 91%<br>(20) | 13%<br>(3) | 87%<br>(20) | 1 | 18 |
| Total | 171 | 24 | 147 | 15 | 147 | 6 | 155 |

## Discussion

Environmentally acquired diseases involve complex interactions between pathogen, host and the environment. Each of these factors play a role in the outcome of infectious diseases. For Valley fever, individuals become exposed to fungal propagules that originate in arid desert soils. Alarmingly, the recognized soil habitat of *Coccidioides spp*. is actively expanding to regions as far north as Washington state and predicted to continue in response to climate variability (55, 57, 58). In the southwestern United States where the desert environment supports fungal growth, exposure is likely inevitable for long term residents. This residential population is steadily increasing, often at a faster rate than the rest of the country combined, which results in an increase in naïve hosts (73). Indeed, these fungal infections are on the rise in the southwestern United States with most cases occurring in California and Arizona (55, 57, 58, 62). The number of reported cases in Arizona have been increasing, and here we focus on the northern counties of the state which are following this trend. In order to create a holistic view of coccidioidomycosis in Northern Arizona, we investigated the patterns of documented susceptible hosts, population structure of the local fungal isolates and the likelihood of the environment supporting the pathogens existence.

In Arizona it is mandatory to report confirmed cases of Valley fever, which allowed us to investigate current reported cases in comparison to historical trends. For each northern Arizona county, the number of total cases and cases per 100,000 population has trended up each year (supplemental tables 3 and 4). In Yavapai, Mohave and Apache counties, the number of cases per 100,000 doubled by 2019 in comparison to historical averages from 2012-2016 (Figure 1, supplemental table 3). Whether these infections are the result of travel to endemic regions of southern Arizona is unknown. Although the results of our soil survey suggest the pathogen may already be present in the region, travel between southern and northern Arizona is common and the more likely explanation (Table 2). We anticipated that regardless of the region of infection, host factors that lead to susceptibility would be similar. As suspected, individuals that test positive for coccidioidomycosis in northern Arizona counties share similar sex and age demographics as compared to state data (Figure 2A, supplemental table 5). Statewide, the ratio of male to females with a confirmed case of Valley fever is typically 50/50 with slight variation (48/52 in 2018) between each year (60). Individuals ages 45-64 are the most commonly diagnosed with Valley fever in Arizona, in the northern Arizona counties similar observations occurred. Because these observations are analogous in northern counties, physicians in the area should consider increased screening for Valley fever, especially among individuals age 45-64 who display symptoms of respiratory infection.

In this study, we demonstrate that thirty-eight cases of Valley fever were treated at the regional medical center during an 18-month period. We set out to compare these northern Arizona in-patients to the greater Arizona statewide Valley fever populations to determine if any other factors were contributing to disease and again found similar trends in regard to age and sex (Table 1, Figure 2). Racial category is a crucial but often missing factor we identified during our study. In total, 57.8% (22/38) of the patients identified as AI/AN (Table 2). Sixty percent is a value higher than expected given that NAH’s typical patient population is 35% AI/AN (Table 1). We acknowledge that many socioeconomic factors such as access to healthy food, income level and quality healthcare can play a role in disease response (74-76). Untangling the complexity of racism and health disparities is a long-standing challenge in epidemiology studies (77). Importantly, overwhelming evidence suggest race as it relates to disease susceptibility is a proxy for a number of underlying factors and that socioeconomic status (SES) rather than racial variation influences health outcomes (29, 77, 78). Nevertheless, in Arizona, over 70% of documented statewide cases do not include race/ethnicity in the disease report. Therefore, identifying an expected distribution of Valley fever cases across racial categories is inaccurate with currently available data (Table 2) (60). The racial diversity in northern Arizona is quite different than the southern parts of the state, specifically in regard to Native populations (79, 80). In total American Indian/Alaskan Natives (AI/AN) make up only 4% of the Arizona population. In contrast, large areas of tribal land exist in Coconino, Navajo and Apache counties hence the percent of tribal members are higher than any other counties in the state (Figure 1 and 2). Therefore, compared to other races and the total population in each county the percentage of AI/AN individuals documented in Coconino county is 26%, in Navajo county is 44% and Apache county is 73%. In contrast, tribal area is smaller in Mohave and Yavapai counties, and therefore the percent of AI/AN in respect to the total populations is 2.2% and 1.6% respectively (supplemental table 6). Research on the susceptibility of native populations to develop severe Valley fever has always suggested concern. In 1974, it was reported that southwestern tribal members had three to five times higher morbidity and mortality rates, compared to whites located in or near the same region, in response to Valley fever (81). A follow up paper in 1985 focused on a decreased dissemination in southern tribes (from 8.9-3.9%) however, elevated mortality rates of native patients compared to that of white populations in the same region still remained (82). More recently, a substantial proportion of Valley fever samples collected in New Mexico were derived from Native American patients (59). At a similar time, the Center for Disease Control (CDC) identified that native populations suffered the highest rate of coccidioidomycosis associated hospitalization out of any other race nationwide (27). The discussion emphasized that increased dissemination and mortality rates of coccidioidomycosis in part were due to delayed diagnosis, which further supports that access to quality healthcare or other SES factors influence disease outcomes(27, 74, 77, 83).

Delays in diagnosis are particularly problematic in symptomatic Valley fever cases. Rapid and accurate diagnosis allows clinicians to monitor disease progression, and determine the appropriate intervention strategies. In contrast, severe disease or dissemination can occur when symptomatic disease is left untreated (25). Our chart review suggests that many of the NAH patients had co-morbidities such as HIV, cancer, or diabetes, which are known to increase disease susceptibility. However, 23% of the patients did not have any immune suppression conditions other than Valley fever, reminding us that this disease can be damaging to otherwise healthy individuals. Of the hospitalized patients 19% had fungal infections disseminate beyond the respiratory system. These data on dissemination are beyond the 1-5% estimate that is expected for Valley fever (25, 32). Interestingly, six or 85% of these disseminated cases identified as AI/AN. This again suggests that tribal members may be at increased risk for dissemination compared to other races, as proposed previously (27, 81). Of note, the CDC report did suggest that the high level of coccidioidomycosis could be due the high proportion of individuals that live in the endemic region (southwestern US). Our study is focused on northern Arizona which includes a large proportion of tribal land (which extends into CO, NM and UT). While this area is in/near the endemic zone, it is not an area traditionally considered to be high risk for acquiring the disease (Figure 1 and 2). The risk may be higher than originally thought based on the increasing county Valley fever case counts per 100,000 population and hospitalizations reported in northern Arizona. Previously, health disparities for AI/AN have been documented for other diseases (83, 84). If NAH is treating a higher proportion of AI/AN with Valley fever than expected, these findings are consistent with calculations performed by the CDC and follow up studies should be designed to address this observation, while considering race and SES factors (27, 78). Whether the risk is due to racism, discrimination, socioeconomic variables, genetic factors, and/or the presence of environmental pathogens, it should be more formally investigated. Our report indicates that a general increased awareness among hospitals, physicians and local populations in Northern Arizona is needed to quickly identify and treat *Coccidioides* infections.

Working with *Coccidioides* spp. requires biosafety level 3 (BSL-3) containment and therefore culturing is not the primary method of diagnosis for most clinical laboratories, limiting the availability of clinical isolates. The seven isolates obtained at NAH represent a unique opportunity to investigate the pathogen component of infections in the region. The *Coccidioides* genus is comprised of two genetically distinct species, each with geographically defined subpopulations (38, 40). *C. immitis* found in California and Washington and *C. posadasii* found in Arizona, Texas, Mexico and Latin America (38, 40-42, 54, 55). Interestingly, a recent publication documented *Coccidioides* isolates from New Mexico patients represent both *C. immitis* and *C. posadasii* infections, it is likely that *C. immitis* infections are travel related (59). Because geographic origin of the fungus can be determined by genetics, we compared the NAH isolates to previously published data sets (40-42). We identified that the newly obtained clinical isolates from Coconino county are *C. posadasii* with genetic proximity to other Arizona isolates that originate from southern AZ counties such as Maricopa and Pima. In contrast, the new clinical isolates do not group with Tex/Mex/SA or Caribbean *C. posadasii* populations. While at the time of sample collection the patients in our study resided in northern Arizona counties, travel/previous residency information was sparse. One patient reported previously living in California and another traveled to northern Mexico, but dates were not provided. These findings demonstrate that these particular NAH patients were infected with isolates from the different Arizona genotypes rather than a unique genetic variant in northern Arizona.

Northern Arizona’s environment is not generally considered preferred habitat for *Coccidioides*. This has been assumed based on the lower number of cases in the area, which, when they are diagnosed, are attributed to travel or previous residency in highly endemic southern regions of the state. Here we provide evidence of *Coccidioides posadasii* DNA in the soil in all of the northern Arizona counties we tested. Interestingly, both CDx and CocciENV assays target alleles that are unique to the genus (91, 96) respectively. Our results demonstrate the presence of *Coccidioides* DNA in a wide range of northern Arizona locations, six of which were positive using both assays, and thus we would expect that cases would be higher across the region. A reasonable explanation would be the presence of a less virulent or novel species in the region, which may not cause severe disease in humans but still be captured by the molecular assays. Reduction in virulence due to evolutionary tradeoff adaptations relate to temperature, precipitation, competition and biodiversity as observed in other species (85, 86). Alternatively, wind dispersed *Coccidioides* arthroconidia from southern endemic regions may deposit conidia, but these have only recently established in response to climate variability as predicted (58). This could be an alternate explanation for the local clinical isolates grouping with southern Arizona populations. The ability of arthroconidia to travel this distance and successfully propagate is unknown and difficult to model. Models predict an expansion of suitable habitat, which includes Northern Arizona, will soon be hospitable to *Coccidioides* in response to climate change. How this will impact already present novel species or wind dispersed disease causing strains is unknown.

## Summary

Here in we discuss the complex interactions between the environmentally associated fungal pathogen *Coccidioides*, which among susceptible human populations is causing the disease coccidioidomycosis in northern Arizona. Our work suggests that the incidence of diagnosed cases is rising in counties located in northern Arizona, some of which are severe enough to require hospitalization. Individuals with confirmed cases demonstrated similar demographics as the rest of the state in regard to sex and age. We observed that 57% of hospitalized patients were of AI/AN heritage, however we are unable to determine if this represents an increase risk without reliable statewide data on race/ethnicity of cases. Increased reporting of patient race statewide is needed to identify disease trends specific to demographics represented in the southwest. Lastly, we identified that the available clinical isolates from the hospital are genetically related to Southern Arizona *C. posadasii* isolates and do not originate from other populations found in Texas, Mexico or California. Our soil survey data suggests that the pathogen is detectable in Northern Arizona soils, which has never previously been recognized. This indicates the region supports the existence of *Coccidioides* species in the environment. Collectively, our work describes the Valley fever disease triangle which considers the interplay between host, pathogen and environment in Northern Arizona. Until disease prevention is a feasible option, we anticipate that incidence of Valley fever will increase in the area. Therefore, enhanced awareness and screening for the disease is vital to the communities of Northern Arizona.

## Study Limitations

To our knowledge this study is the first attempt to consider the interaction of host, pathogen and environment which contribute to Valley fever in northern Arizona. Therefore, results and discussion presented here are an estimate, with need for further data collection. Our goal was to document Valley fever trends in northern Arizona with data at hand. We present Valley fever cases and cases per 100,000 to account for reported cases and population fluctuation. However, we acknowledge that the number of Valley fever tests administered (positive and negative) can impact these numbers. We did not have access to these data, and thus this is a limitation of our interpretation. Additionally, individuals with Valley fever must seek medical care and be tested to be included in the reported case counts. Subsequently, our county data and in-patient hospital data do not reflect asymptomatic cases, individuals without access to medical care or misdiagnosis. Ideally, the retrospective chart review would encompass a longer time span, however due to the current pandemic we chose to present the current data rather than request further hospital resources. As previously mentioned, culturing is not the primary determinate for Valley fever diagnosis, therefore we had limited access to clinical isolates. While we were able to determine the genetic relationship of these isolates described above, we cannot make inferences about the genetic or geographic relationship of infections where clinical isolates were not collected. Lastly, the PCR methods utilized in this study detect *Coccidioides* DNA, which does not confirm the presence of live infectious organisms, but rather suggests it.

## Data Availability

Raw reads used in this study have been deposited under accession number PRJNA722304.

## Acknowledgments

Appreciation to note.louis designs for map/figure creation (fig1,fig 4).

