## supplemental table 3 for "Disease Triangle Dynamics of Coccidioidomycosis in Northern Arizona"

Supplemental Table 3 Reported cases/100,000 population

| **County** | **2017** | **2018** | **2019** | **Historical Average 2012-2016** |
| --- | --- | --- | --- | --- |
| Apache | 41.8 | 28.6 | 57.0 | 22.7 |
| Cochise | 23.7 | 34.5 | 63.5 | 38.8 |
| Coconino | 21.7 | 31.6 | 36.9 | 24.8 |
| Gila | 112.4 | 83.7 | 135.1 | 75.9 |
| Graham | 46.8 | 47.2* | 41.1 | 51.2 |
| Greenlee | 28.9 | 9.5* | 10.5 | 27.5 |
| La Paz | 80.6 | 64* | 104.2 | 76.9 |
| Maricopa | 112.9 | 128 | 162.5 | 140.0 |
| Mohave | 30.9 | 39.9 | 66.4 | 32.4 |
| Navajo | 40.8 | 37.3 | 61.3 | 36.8 |
| Pima | 97.8 | 94.6 | 136.5 | 110.5 |
| Pinal | 113.8 | 129.1 | 201.0 | 139.8 |
| Santa Cruz | 32.0 | 38.2 | 53.8 | 30.7 |
| Yavapai | 25.8 | 31 | 51.9 | 23.5 |
| Yuma | 12.2 | 12 | 30.4 | 13.3 |

*Less than 20 cases so number is unreliable

Retrieved from Arizona Department of Health website.
