## supplemental table 4 for "Disease Triangle Dynamics of Coccidioidomycosis in Northern Arizona"

Supplemental Table 4 Reported cases

| **County** | **2017** | **2018** | **2019** |
| --- | --- | --- | --- |
| Maricopa | 4,932 | 5,495 | 7,291 |
| Pima | 1,022 | 978 | 1,430 |
| Pinal | 518 | 569 | 930 |
| Mohave | 67 | 82 | 141 |
| Gila | 62 | 46 | 73 |
| Yavapai | 60 | 71 | 122 |
| Navajo | 46 | 42 | 68 |
| Coconino | 32 | 46 | 53 |
| Cochise | 31 | 45 | 80 |
| Apache | 30 | 21 | 41 |
| Yuma | 28 | 27 | 65 |
| La Paz | 19 | 14 | 22 |
| Graham | 18 | 18 | 16 |
| Santa Cruz | 17 | 20 | 25 |
| Greenlee | 3 | 1 | 1 |

Retrieved from Arizona Department of Health website.
