## supplemental table 5 for "Disease Triangle Dynamics of Coccidioidomycosis in Northern Arizona"

Supplemental table 5. Observed incidence of gender and age for Valley fever patients in northern Arizona counties

| Total population and confirmed Valley fever cases for state*, county*, hospital demographics** | | | | | | |
| --- | --- | --- | --- | --- | --- | --- |
|  | Sex | | Age | | | |
|  | M | F | <25 | 25-44 | 45-64 | 65-84 |
| Arizona | 48%  (3,572) | 52%  (3,885) | 825 | 1,784 | 2,425 | 2,183 |
| Mohave | 56%  (48) | 44%  (37) | 5 | 9 | 29 | 42 |
| Yavapai | 51%  (36) | 49%  (35) | 4 | 12 | 19 | 36 |
| Coconino | 63%  (29) | 37%  17 | 6 | 16 | 14 | 11 |
| Navajo | 48%  (20) | 52%  (22) | 2 | 14 | 14 | 14 |
| Apache | 57%  (12) | 43%  (9) | 1 | 4 | 11 | 5 |
| Northern AZ Healthcare | 60.5%  (23) | 39.4%  (15) | 3 | 12 | 16 | 6 |

*Arizona Department of Health 2018 annual report

** Northern Arizona Healthcare, Flagstaff Location 2017, 2018
