## supplemental table 6 for "Disease Triangle Dynamics of Coccidioidomycosis in Northern Arizona"

| POPULATION BY FIVE-YEAR AGE GROUPS, COUNTY, GENDER, AND RACE/ETHNICITY, ARIZONA, 2019 | | | | |
| --- | --- | --- | --- | --- |
|  | | | Total | Percent |
| Arizona | All groups | Total | 7,189,020 | 100.00 |
|  |  | Male | 3,574,659 | 49.72 |
|  |  | Female | 3,614,361 | 50.28 |
|  | White non-Hispanic | Total | 3,981,049 | 55.38 |
|  |  | Male | 1,968,918 | 27.39 |
|  |  | Female | 2,012,131 | 27.99 |
|  | Hispanic or Latino | Total | 2,279,253 | 31.7 |
|  |  | Male | 1,146,980 | 15.95 |
|  |  | Female | 1,132,273 | 15.75 |
|  | Black or African American | Total | 352,121 | 4.898 |
|  |  | Male | 182,011 | 2.532 |
|  |  | Female | 170,110 | 2.366 |
|  | American Indian or Alaska Native | Total | 299,123 | 4.161 |
|  |  | Male | 145,181 | 2.019 |
|  |  | Female | 153,942 | 2.141 |
|  | Asian or Pacific Islander | Total | 277,474 | 3.86 |
|  |  | Male | 131,569 | 1.83 |
|  |  | Female | 145,905 | 2.03 |

| POPULATION BY FIVE-YEAR AGE GROUPS, APACHE COUNTY, GENDER, AND RACE/ETHNICITY, ARIZONA, 2019 | | | | |
| --- | --- | --- | --- | --- |
|  | | | Total | Percent |
| Apache | Total | Total | 71,808 | 0.999 |
|  |  | Male | 35,307 | 49.17 |
|  |  | Female | 36,501 | 50.83 |
|  | White non-Hispanic | Total | 13,313 | 18.54 |
|  | Hispanic or Latino | Total | 4,589 | 6.391 |
|  | Black or African American | Total | 640 | 0.891 |
|  | American Indian or Alaska Native | Total | 52,923 | **73.7** |
|  | Asian or Pacific Islander | Total | 343 | 0.478 |

| POPULATION BY FIVE-YEAR AGE GROUPS, COCONINO COUNTY, GENDER, AND RACE/ETHNICITY, ARIZONA, 2019 | | | | |
| --- | --- | --- | --- | --- |
|  | | | Total | Percent |
| Coconino | Total | Total | 147,275 | 2.049 |
|  |  | Male | 72,778 | 49.42 |
|  |  | Female | 74,497 | 50.58 |
|  | White non-Hispanic | Total | 81,209 | 55.14 |
|  | Hispanic or Latino | Total | 21,023 | 14.27 |
|  | Black or African American | Total | 2,496 | 1.695 |
|  | American Indian or Alaska Native | Total | 39,187 | 26.61 |
|  | Asian or Pacific Islander | Total | 3,360 | 2.281 |

| POPULATION BY FIVE-YEAR AGE GROUPS, NAVAJO COUNTY, GENDER, AND RACE/ETHNICITY, ARIZONA, 2019 | | | | |
| --- | --- | --- | --- | --- |
|  | | | Total | Percent |
| Navajo | Total | Total | 112,825 | 1.56941 |
|  |  | Male | 56,472 | 50.0527 |
|  |  | Female | 56,353 | 49.9473 |
|  | White non-Hispanic | Total | 47,698 | 42.2761 |
|  | Hispanic or Latino | Total | 12,928 | 11.4585 |
|  | Black or African American | Total | 1,348 | 1.19477 |
|  | American Indian or Alaska Native | Total | 49,995 | 44.312 |
|  | Asian or Pacific Islander | Total | 856 | 0.7587 |

| POPULATION BY FIVE-YEAR AGE GROUPS, MOHAVE COUNTY, GENDER, AND RACE/ETHNICITY, ARIZONA, 2019 | | | | |
| --- | --- | --- | --- | --- |
|  | | | Total | Percent |
| Mohave | Total | Total | 216,985 |  |
|  |  | Male | 109,728 | 50.56939 |
|  |  | Female | 107,257 | 49.43061 |
|  | White non-  Hispanic | Total | 169,143 | 77.95147 |
|  | Hispanic or  Latino | Total | 36,533 | 16.83665 |
|  | Black or  African  American | Total | 3,028 | 1.395488 |
|  | American  Indian or  Alaska Native | Total | 4,941 | 2.277116 |
|  | Asian or Pacific Islander | Total | 3,340 | 1.539277 |

| POPULATION BY FIVE-YEAR AGE GROUPS, YAVAPAI COUNTY, GENDER, AND RACE/ETHNICITY, ARIZONA, 2019 | | | | |
| --- | --- | --- | --- | --- |
|  | | | Total | Percent |
| Yavapai | Total | Total | 232,386 | 3.232513 |
|  |  | Male | 113,515 | 48.84761 |
|  |  | Female | 118,871 | 51.15239 |
|  | White non-  Hispanic | Total | 189,145 | 81.3926 |
|  | Hispanic or  Latino | Total | 34,115 | 14.68032 |
|  |  | Male | 17,529 | 7.543053 |
|  |  | Female | 16,586 | 7.137263 |
|  | Black or  African American | Total | 2,279 | 0.980696 |
|  | American  Indian or  Alaska Native | Total | 3,800 | 1.63521 |
|  | Asian or  Pacific Islander | Total | 3,047 | 1.311181 |
